# Drinking and smoking polygenic risk is associated with neurodevelopmental outcomes of children and young adults independently of psychopathology and substance use

**DOI:** 10.1101/2020.10.22.20217596

**Authors:** Flavio De Angelis, Frank R. Wendt, Gita A. Pathak, Daniel S. Tylee, Aranyak Goswami, Joel Gelernter, Renato Polimanti

**Author notes:** **Correspondence to** Renato Polimanti, PhD; VA CT 116A2, 950 Campbell Avenue, West Haven, CT, 06516, USA. **Conflict of Interest:** Dr. Polimanti is paid for his editorial work on the journal Complex Psychiatry. The other authors declare no competing interests.

## Abstract

**Background:** Alcohol drinking and tobacco smoking are hazardous behaviors associated with a wide range of adverse health outcomes, including many mental and physical disorders.

**Methods:** To investigate the pleiotropic mechanisms linking these traits to cognitive and behavioral development, we explored the association of polygenic risk scores (PRS) related to drinks per week (DPW), age of smoking initiation (ASI), smoking initiation (SI), cigarettes per day (CPD), and smoking cessation (SC) with 433 neurodevelopmental features in 4,498 children and young adults of European ancestry from the Philadelphia neurodevelopmental cohort (PNC). This sample was not enriched for specific psychiatric traits, but 21% of the PNC participants endorsed substance use.

**Results:** After applying a false discovery rate multiple testing correction accounting for the number of PRS and traits tested, we identified 36 associations related to psychotic symptoms, emotion and age recognition social competencies, verbal reasoning, anxiety-related traits, parents’ education, and substance use. These associations were independent of the genetic correlations among the alcohol-drinking and tobacco-smoking traits and those with cognitive performance, educational attainment, risk-taking behaviors, and psychopathology. The removal of participants endorsing substance use did not affect the associations of each PRS with neurodevelopmental traits identified as significant in the discovery analyses. Gene-ontology enrichment analyses identified several neurobiological processes underlying mechanisms of the PRS associations we report. These were mainly related to brain connectivity.

**Conclusions:** We provide novel insights into the genetic overlap of smoking and drinking behaviors with neurodevelopment in children and young adults, highlighting their independence from psychopathology and other substance use.

## Introduction

Alcohol drinking and tobacco smoking may result in direct or indirect health concerns. Psychoactive compounds such as ethanol and nicotine act primarily on mental processes and therefore change mood, feelings, and behavior (WHO, 2010), but they are also related to many negative health outcomes (WHO, 2012 and 2018).Drinking and smoking representing two of the three leading preventable causes of death in the US (NSDUH 2018). Understanding the molecular and behavioral processes underlying the predisposition to alcohol drinking and tobacco smoking could lead to better prevention strategies to reduce the harmful consequences associated with these behaviors. Large-scale genome-wide association studies (GWAS) of traits related to alcohol drinking and tobacco smoking demonstrated that the predisposition to these complex behavioral traits is highly polygenic (i.e., thousands of variants with small effects) (Clarke et al., 2017; Jorgenson et al., 2017; Gelernter et al., 2018; Kranzler et al., 2019; Erzurumluoglu et al., 2019, Matoba et al., 2019, Nana et al., 2019). To date, GSCAN (GWAS & Sequencing Consortium of Alcohol and Nicotine) has completed the largest genome-wide meta-analysis across multiple drinking and smoking behaviors short of dependence on either of these substances, analyzing up to 1.2 million individuals (Liu et al., 2019). GSCAN investigated one alcohol-drinking phenotype (drinks per week, DPW) and four tobacco-smoking phenotypes. These included cigarettes per day (CPD, average number of cigarettes smoked per day), smoking initiation (SI, smoker versus non-smoker), smoking cessation (SC, current versus former smoker), and age of smoking initiation (ASI, age at which an individual started smoking regularly). While ASI is negatively genetically correlated with all the other traits (from rg=-0.10 with respect to DPW to rg=-0.71 for SI), DPW and the other smoking phenotypes share positive genetic correlations ranging from rg=0.07 (CPD vs. DPW) to rg=0.42 (SC vs. CPD). These traits also showed a broad spectrum of genetic correlations including behavioral traits (e.g., risk tolerance and neuroticism), psychiatric disorder (e.g., major depressive disorder and schizophrenia), and physical health outcomes (e.g., obesity and coronary artery disease) (Liu et al., 2019). Due to the large effects of tobacco and alcohol on human health, it is challenging and important to distinguish whether the genetic correlations observed are due to the consequences of alcohol drinking and tobacco smoking or to the genetic etiology shared between these traits and other complex phenotypes. To work towards dissecting the pleiotropic mechanisms related to alcohol drinking and tobacco smoking, we investigated polygenetic risk scores derived from GSCAN genome-wide association data with respect to neurodevelopmental features assessed in the Philadelphia Neurodevelopmental Cohort (PNC). This is a population-based sample including children and young adults recruited at the Children’s Hospital of Philadelphia (CHOP). Each PNC participant was assessed neuropsychiatrically with a structured interview and completed a computerized neurocognitive battery (CNB). Due to the limited alcohol and tobacco use in the young CHOP participants, we investigated the association of genetic liability to alcohol drinking and tobacco smoking with neurodevelopmental traits, independently from the potential effects of psychoactive compounds such as nicotine and alcohol. In other words, this study design permits detection of association between smoking and drinking trait polygenic risk and neurodevelopment, without confounding effects of substance use, as would likely be present in an adult cohort. Additionally, we also verified that the associations observed are not due to the genetic overlap across the GSCAN phenotypes or to other genetically correlated psychiatric and behavioral traits including psychopathology, risk tolerance, educational attainment, and socioeconomic status.

## Materials and Methods

### Philadelphia Neurodevelopmental Cohort

Neuropsychiatric and genotype data for PNC participants were obtained, after authorized access, from the National Center for Biotechnology Information (NCBI) database of Genotypes and Phenotypes (dbGaP; available at http://www.ncbi.nlm.nih.gov/gap) through dbGaP accession number phs000607.v3.p2 (Neurodevelopmental Genomics: Trajectories of Complex Phenotypes). The PNC is a population-based sample including of more than 9,500 individuals aged 8-21 years not enriched for any epidemiologically ascertained specific disorder, behavior, or trait (Satterthwaite et al., 2016). The PNC participants were randomly recruited after stratification by sex, age, and ethnicity. Individuals recruited presented diverse medical conditions that were summarized in a medical rating, ranging from minor issues to potentially life-threatening health concerns. Each participant was assessed for neuropsychiatric traits with a structured interview and completed a computerized neurocognitive battery (CNB) (Moore et al., 2015). The structured interview included a panel of questions related to demographics, the timeline of life events, education, medical history, psychopathological assessment, and a global assessment of cognitive and executive functioning. The CNB consisted of 14 tests assessing executive-control, episodic memory, complex cognition, social cognition, and sensorimotor speed. Young adults (ages 18-21) were interviewed, while children aged 8-10, and 11-17 were submitted to collateral assessment with a parent. To account for the overall medical conditions and the assessment strategy, we included medical rating and type of interview among the covariates of the regression models.

Our analysis was restricted to PNC participants of European descent due to the lack of availability of large-scale GWAS data for other populations and known biases of cross-ancestry PRS analysis (Martin et al., 2017). Considering these inclusion criteria, we investigated 433 neurodevelopmental features (Supplementary Table 1) in 4,498 children and young adults of European descent. Data quality control was performed as detailed in Wendt et al., 2019. Using Plink v1.9 (Chang et al., 2015), we removed variants with call rate < 0.95 and HWE p-values<1×10^−6^. Our study only investigated unrelated subjects considering a phi-hat threshold of 0.2. European ancestry was confirmed via principle component analysis using linkage-disequilibrium independent SNPs shared across all PNC bead chips and the 1000 Genomes Project reference panel for European populations (N=503). SHAPEIT and IMPUTE2 (O’ Connell et al., 2014; Howie et al., 2009) were employed in pre-phasing and imputation with the 1000 Genomes Project Phase 3 set as a reference.

### Genome-wide association data

Genome-wide association data for drinking and smoking traits were derived from GSCAN (Liu et al., 2019) and accessed via the Data Repository of the University of Minnesota (available at https://conservancy.umn.edu/handle/11299/201564). GSCAN GWAS included only individuals of European descent. GWAS data were generated in each study included in the GSCAN GWAS using RVTEST (Zhan et al., 2016), accounting for the family-based studies and unrelated samples (Liu et al., 2019). GSCAN investigated five traits, one related to alcohol drinking and four related to tobacco smoking. DPW (N=941,280) was defined based on the average number of alcoholic drinks a participant reported consuming in a week. SI (N=1,232,091) is a binary trait considering regular smokers as cases and non-smokers as controls, while ASI (N=341,427) is a quantitative trait related to the age when an individual started to smoke tobacco-based cigarettes. SC (N=547,219) was defined considering current smokers as cases and former smokers as controls. CPD (N=337,334) was calculated in both current and former smokers by binning the quantitative measure of CPD (bin1=1-5; bin2=6-15; bin3=16-25; bin4=26-35; bin5=36+).

To verify whether the polygenic risk scores (PRS) associations were due to the genetic correlation of alcohol drinking and tobacco smoking with other complex traits, we investigated other large-scale GWAS datasets, including Psychiatric Genomics Consortium cross-disorder (PGC-CD; N=438,997, Cross-Disorder Group of the Psychiatric Genomics Consortium, 2019), Social Science Genetic Association Consortium (SSGAC) cognitive performance (N=257,828, Lee et al., 2018), SSGAC educational attainment (N=766,345, Lee et al., 2018), SSGAC general-risk-tolerance (N=466,571, Karlsson-Linner et al., 2019), and household income (N=286,301, Hill et al., 2019) (Supplementary Table 2). The PGC-CD study is a cross-disorder analysis including anorexia nervosa, attention-deficit/hyperactivity disorder, autism spectrum disorder, bipolar disorder, major depression, obsessive-compulsive disorder, schizophrenia, and Tourette syndrome (Cross-Disorder Group of the Psychiatric Genomics Consortium, 2019). We used PGC-CD GWAS data to account for the genetic overlap of alcohol and tobacco use with psychopathology and psychiatric comorbidities. SSGAC educational attainment (EA) was defined by mapping the major educational qualification of each participant to relevant categories from the International Standard Classification of Education (ISCED), then imputing the years-of-education equivalent for each ISCED category, facilitating comparison across different systems (Lee et al., 2018). The SSGAC cognitive performance (CP) data were generated by meta-analyzing data from the COGENT (Cognitive Genomics Consortium) study and the UK Biobank. The COGENT study analyzed a phenotype defined as the first principal component derived from three or more neuropsychological tests (Trampush et al., 2017). In UKB, cognitive performance was defined based on the respondent’s score on a test of verbal-numerical reasoning (Lee et al., 2018). The SSGAC GWAS of general risk tolerance (GR) meta-analyzed cohorts with different assessments capturing an individual’s tendency, preparedness, or willingness to take risks in general (Karlsson Linner et al., 2019). Annual household income (HI) GWAS was conducted in UKB using self-reported pre-tax household income binned to create a five-point scale (bin1<£18,000, bin2=£18,000–£29,999,bin3=£30,000–£51,999,bin4=£52,000–£100,000, bin5>£100,000 (Hill et al., 2019). HI data were analyzed as a proxy of socioeconomic status.

### SNP-based heritability and genetic correlation

SNP-based heritability and genetic correlation for the smoking and drinking traits, and the additional phenotypes were estimated using the Linkage Disequilibrium Score Regression (LDSC) method (Bulik-Sullivan et al., 2015, Finucane et al., 2015). As provided by the LDSC developers (details available at https://github.com/bulik/ldsc), the analysis was conducted considering the HapMap 3 reference panel and pre-computed LD scores based on the 1000 Genomes Project reference data for individuals of European ancestry.

### PRS analysis

PRS based on drinking and smoking GWAS data were investigated with respect to neurodevelopmental traits in the PNC. PRSice v. 2.3.1.c (Choi and O’Reilly, 2019) was used to compute PRS. SNPs were clumped based on 250kb windows, based on clump-r2 threshold = 0.1 and clump-p threshold = 1, respectively. The step size of the threshold was set to 5e-05, and the starting p-value threshold was 5e-08 using an additive model for regression at each threshold. In our analysis, we tested phenotypes that were assessed in ≥ 500 participants. These were grouped in 18 domains: attention deficit disorder, depression, generalized anxiety disorder, outcome of neuropsychiatric tests, mania/hypomania, medical concerns, obsessive compulsive disorder, oppositional defiant disorder, panic disorder, specific phobia, psychosis, post-traumatic stress, general probes, separation anxiety, structured interview for prodromal symptoms, social anxiety, substance use, and other (i.e., phenotypes not classifiable in previous domains). All PRS were covaried for age, sex, the first ten within-ancestry principal components (PCs), medical rating – the PNC code indicating the severity of a patient’s medical condition, and type of interview. False discovery rate (FDR) at 5% was applied to correct the results obtained for multiple testing, accounting for the number of phenotypes and PRS tested. The Manhattan plot related to the phenome-wide association study using the phenotypic binning was generated in R using the ggplot2 package (Wickham, 2009). The individual genetic liabilities to DPW, ASI, SI, CPD, and SC were split into quartiles and regressed against the phenotype of interest using a generalized linear model. For PNC phenotypes significantly associated with the PRS, we also calculated Spearman’s rank-order correlations via R using the rcorr function of the Hmisc library (Alzola and Harrell, 2006). Correlation p-values were adjusted for the number of tests performed using FDR q < 0.05. Finally, we verified whether the significant PRS associations were independent from the genetic correlation among alcohol and tobacco traits and between them and other psychiatric and behavioral traits. The association of each significant PRS with a specific neurodevelopmental feature was tested for the independence of the genetic correlation across the smoking and drinking phenotypes, adding the remaining alcohol and tobacco PRS as additional covariates. Similarly, significant PRS associations were also covaried using PRS related to other psychiatric and behavioral traits (Supplementary Table 2).

### Enrichment analysis

The SNPs used to generate each significant PRS association were analyzed for pathway enrichment using PRSet implemented in PRSice v. 2.3.1.c (Choi and O’Reilly, 2019). A collection of gene sets related to Gene Ontology (GO) terms was downloaded from the Molecular Signatures Database (MSigDB, https://www.gsea-msigdb.org/gsea/msigdb/), and a GTF file containing the genome boundary of the genic regions (Genome build GTCh38.p7) was downloaded from Ensembl repository (Yates et al., 2019) (ftp://ftp.ensembl.org/pub/release-86/gtf/homo_sapiens). REVIGO (Supek et al., 2011) was employed to summarize GO terms by removing redundant items based on Jiang and Conrath semantic distance (Couto et al., 2007) and a similarity degree of 0.5.

## Results

### SNP-based heritability and genetic correlation

The SNP-based heritability of the alcohol-drinking and tobacco-smoking traits ranged from 0.047±0.003 for ASI to 0.072±0.007 for CPD (Supplementary Table 3). LDSC-based correlations were examined among the substance-use phenotypes. DPW showed a positive genetic correlation with each tobacco-smoking trait (Smoking Initiation, SI rg=0.407, P=1.40E-92; Cigarettes per Day, CPD rg=0.083, P=3.93E-03; Smoking Cessation, SC rg=0.108, P=1.02E-03; Figure 1; Supplementary Table 4) with the exception of ASI (Age of Smoking Initiation, rg=-0.160, P=2.73E-07). Similarly, ASI is negatively genetically correlated with the other tobacco-smoking phenotypes (ASI vs. SI, rg=-0.684, P=3.09E-199; ASI vs. CPD, rg=-0.369, P=1.07E-23; ASI vs. SC, rg=-0.291, P=1.71E-12). As we would also verify that the associations observed are not due to the genetic overlap to other genetically correlated psychiatric and behavioral traits, LDSC was used to estimate the SNP-based heritability of these traits (Supplementary Table 2).

**Figure 1:**
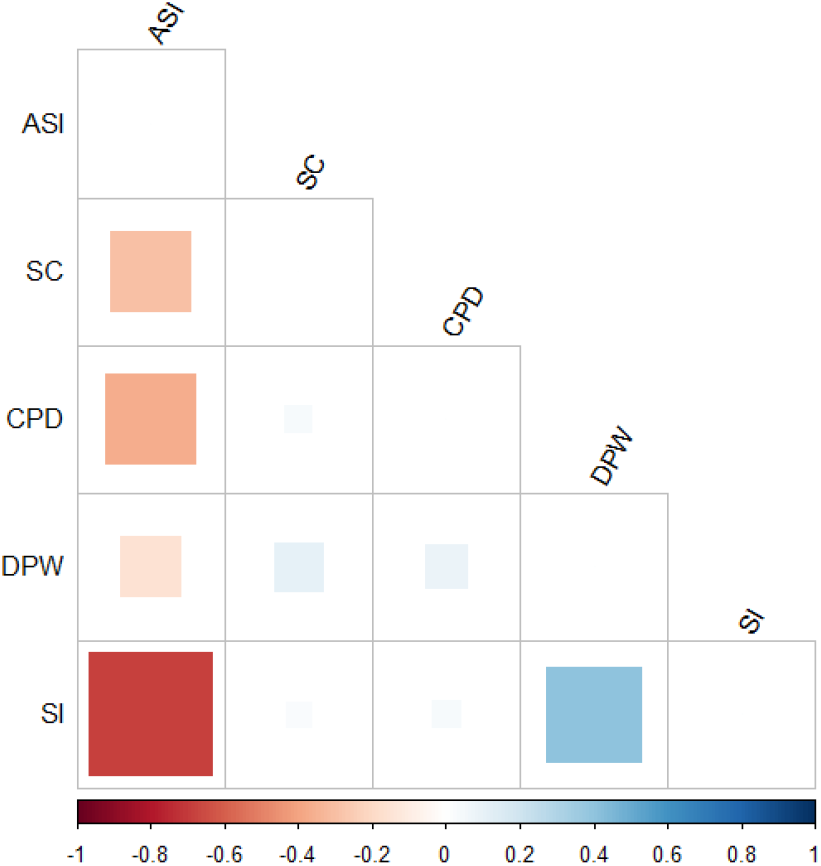
Genetic correlation matrix among smoking and drinking PRS (ASI: Age of Smoking Initiation, CPD: Cigarettes per day, DPW: Drinks per Week, SC: Smoking cessation, SI: Smoking Initiation). The square size is proportional to the magnitude of the correlation. Blank squares relate to not significant correlations (P>0.01).

PGC-CD (Psychiatric Genomics Consortium Cross Disorder) showed a negative genetic correlation with CPD (rg=-0.21, P=2.13E-18), SI (rg=-0.215, P=2.33E-26), DPW (rg=-0.107, P=6.01E-06), and SC (rg=-0.102, P=1.54E-03), while a positive correlation was present with respect to ASI (rg=0.147; P=9.22E-09). CP (Cognitive Performance) was positively correlated with ASI (rg=0.314, P=6.31E-32), while the other smoking PRS showed only a weak negative rg (CP vs. SI rg=-0.172, P=2.34E-22; CP vs. CPD rg=-0.103, P=2.35E-05, and CP vs. SC rg=-0.298, P=2.80E-28).

EA (Educational Attainment) was the most correlated additional behavioral trait respect to ASI (rg=0.599, P=7.08E-167), SC (rg=-0.502, P=4.15E-95), SI (rg=-0.362, P=2.75E-128), and CPD(rg=-0.285, P=8.32E-29). HI (Household Income) followed the same correlation pattern of EA, while risk taking behavior (GR) is the only adjunctive trait negatively related to ASI (rg=-0.228, P=8.46E-13), showing a positive association with the other smoking phenotypes (GR vs. SI rg=0.327, P=1.54E-46; R vs. CPD rg=0.175, P=7.87E-07; GR vs. SC rg=0.180, P=4.22E-07).Finally, GR was also the only additional trait with a moderate positive genetic correlation with DPW (rg=0.286, P=4.26E-30) (Supplementary Table 4).

### Neurodevelopmental Trait Prediction

The genetic liabilities to drinking and smoking behaviors were tested with respect to neuropsychiatric phenotypes assessed in PNC children and young adults. A total of 36 neurodevelopmental traits were significantly associated with alcohol-drinking and tobacco-smoking PRS after accounting for the number of PRS and phenotypes tested (FDR<5%; Table 1; Figure 2, Supplementary Table 5).

**Table 1:**
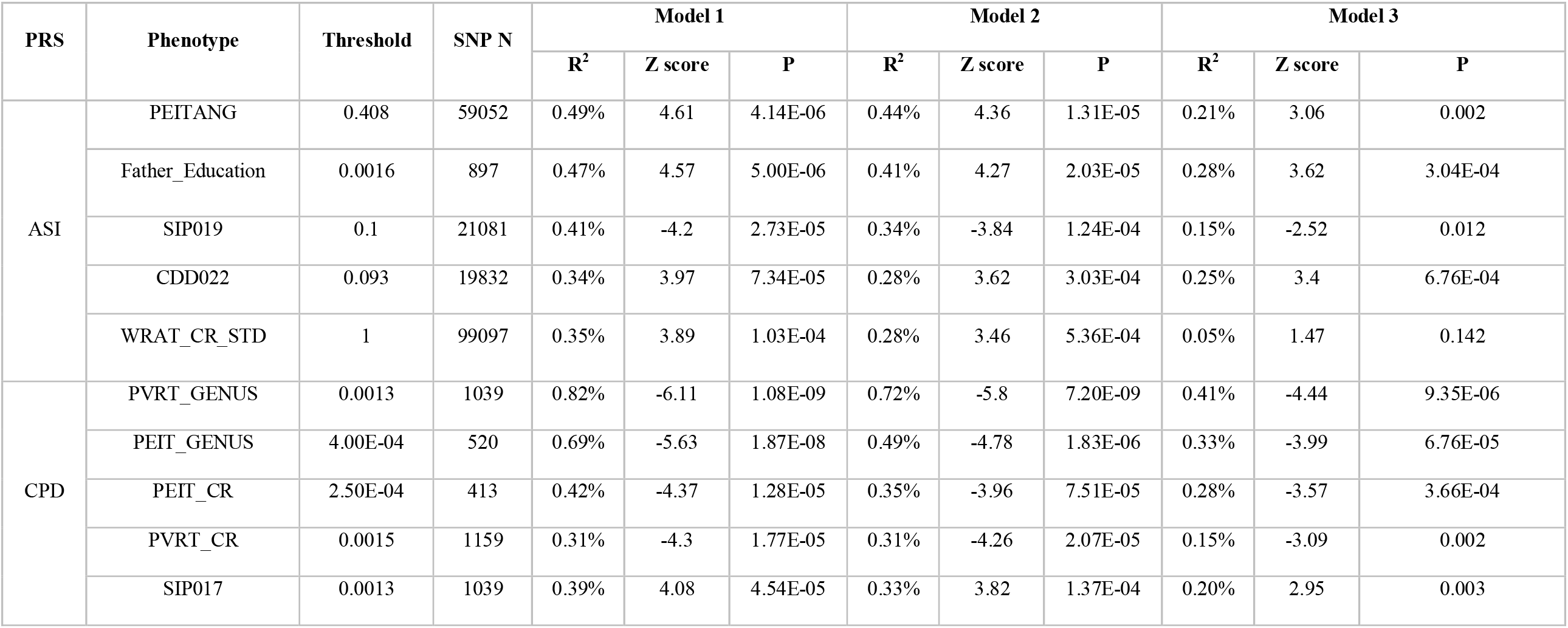
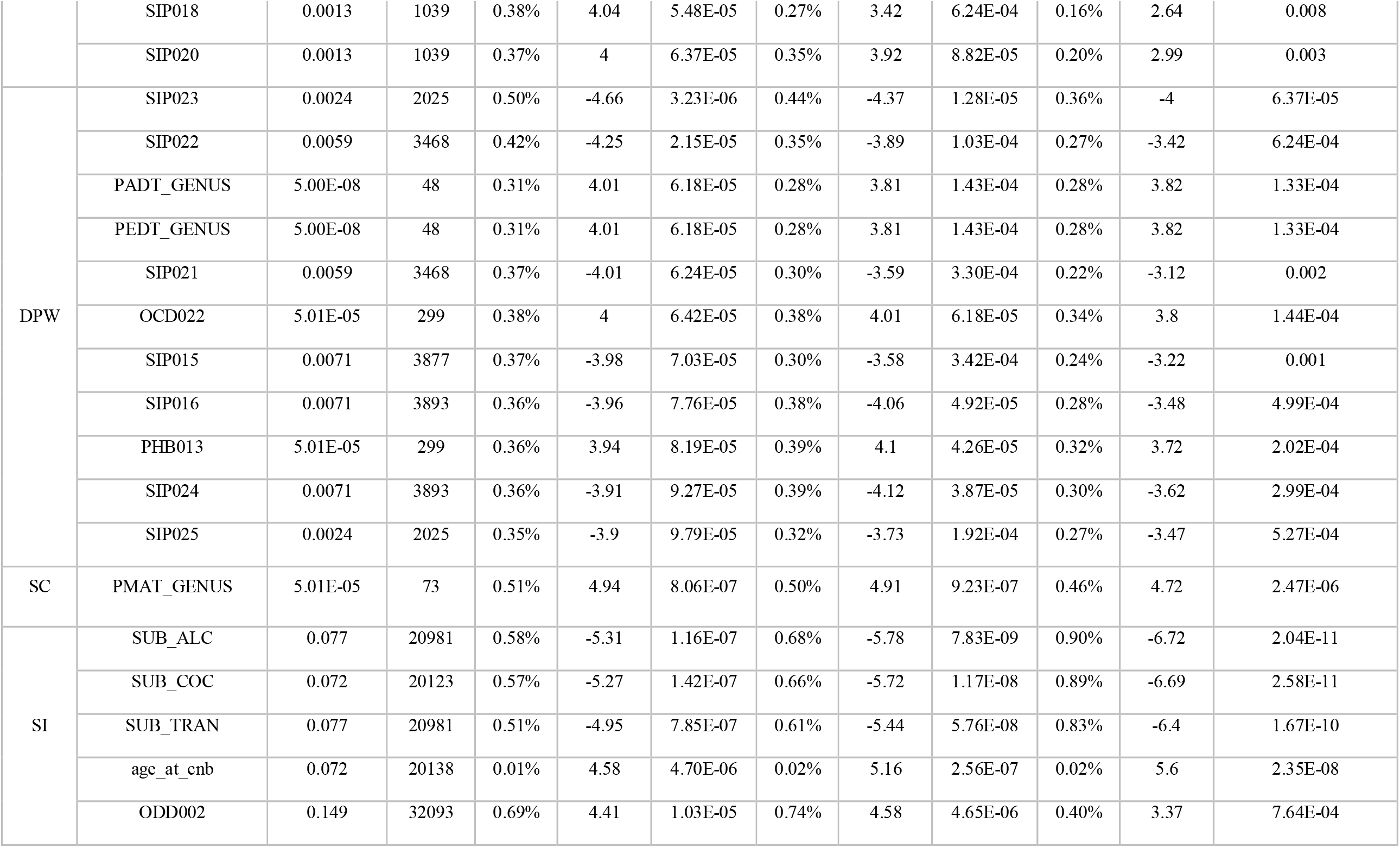
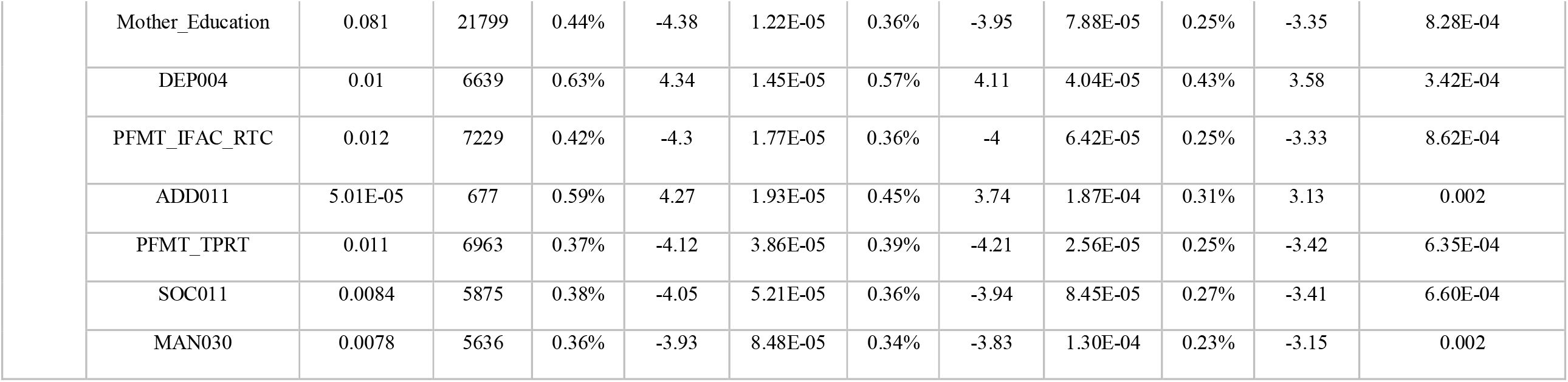
Association of smoking and drinking PRS (ASI: Age of Smoking Initiation, CPD: Cigarettes per day, DPW: Drinks per Week, SC: Smoking cessation, SI: Smoking Initiation) with neurodevelopmental phenotypes surviving the FDR 5% threshold. The legend for each neurodevelopmental code is reported in Supplemental Table 1. Model 1 covariates: 10 principal components (PCs), Sex, Age, Int_Type (Type of Interview), Med_Rate (Medical Rating); Model 2 covariates: 10 PCs, Sex, Age, Int_Type, Med_Rate, other smoking and alcohol PRS; Model 3 covariates: 10 PCs, Sex, Age, Int_Type, Med_Rate, PGC cross-disorder, cognitive performance, educational attainment, general risk tolerance, and household income.

**Figure 2:**
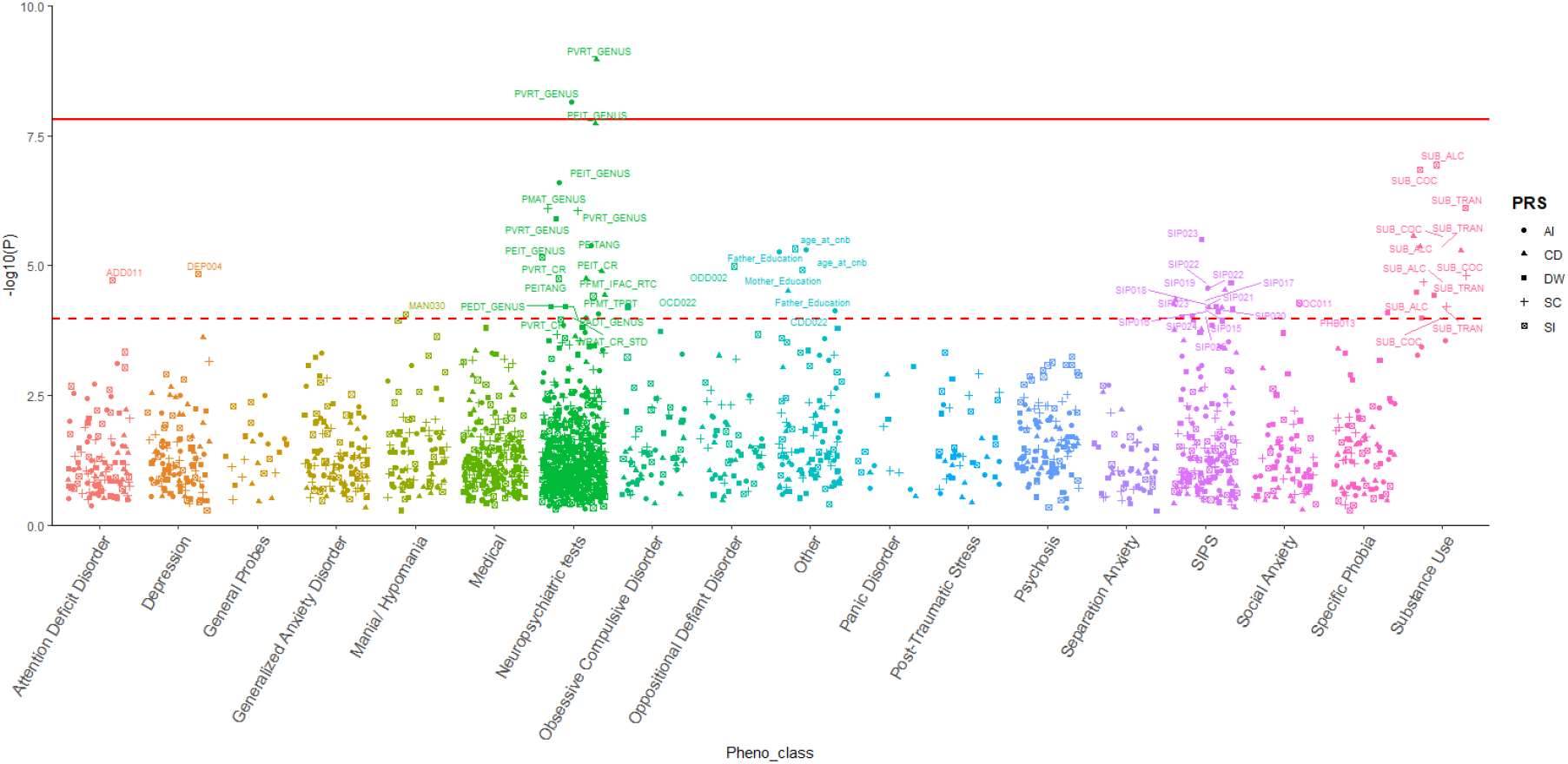
Phenome-wide association study (PheWAS) plot for 433 neurodevelopmental traits and 5 polygenic risk scores (PRS; ASI: Age of Smoking Initiation, CPD: Cigarettes per day, DPW: Drinks per Week, SC: Smoking cessation, SI: Smoking Initiation). The dashed line represents the FDR 5% threshold, while the solid line refers to Bonferroni 5% correction. The legend for each neurodevelopmental code is reported in Supplemental Table 1.

Considering the phenotypic correlation of these 36 neurodevelopmental features (Figure 3; Supplementary Table 6), we observed high correlation among the outcomes derived from the Structured Interview for Prodromal Symptoms (SIPS) (Miller et al., 1999) (Spearman’s ρ>0.87, P<0.0001). Penn computerized individual tests outcomes were also positively correlated to each other: Median Response Time for Correct Responses to Target Faces (PFMT_TPRT) and Median Response Time for Total Correct Test Trial Responses (PFMT_IFAC_RTC) (Spearman’s ρ=0.87, P<0.0001) for Penn Face Memory Test; Penn Verbal Reasoning Test Genus (PVRT_GENUS) and Penn Matrix Reasoning Test Genus (PMAT_GENUS) (Spearman’s ρ=0.61, P<0.0001) for verbal and reasoning tests; Penn Age Differentiation Test (PADT_GENUS) and Penn Emotion Differentiation Test (PEDT_GENUS) (Spearman’s ρ=1, P<0.0001) for age and emotion recognition. Alcohol, cocaine, and tranquilizer use were also highly correlated (Spearman’s ρ>0.98, P<0.0001).

**Figure 3:**
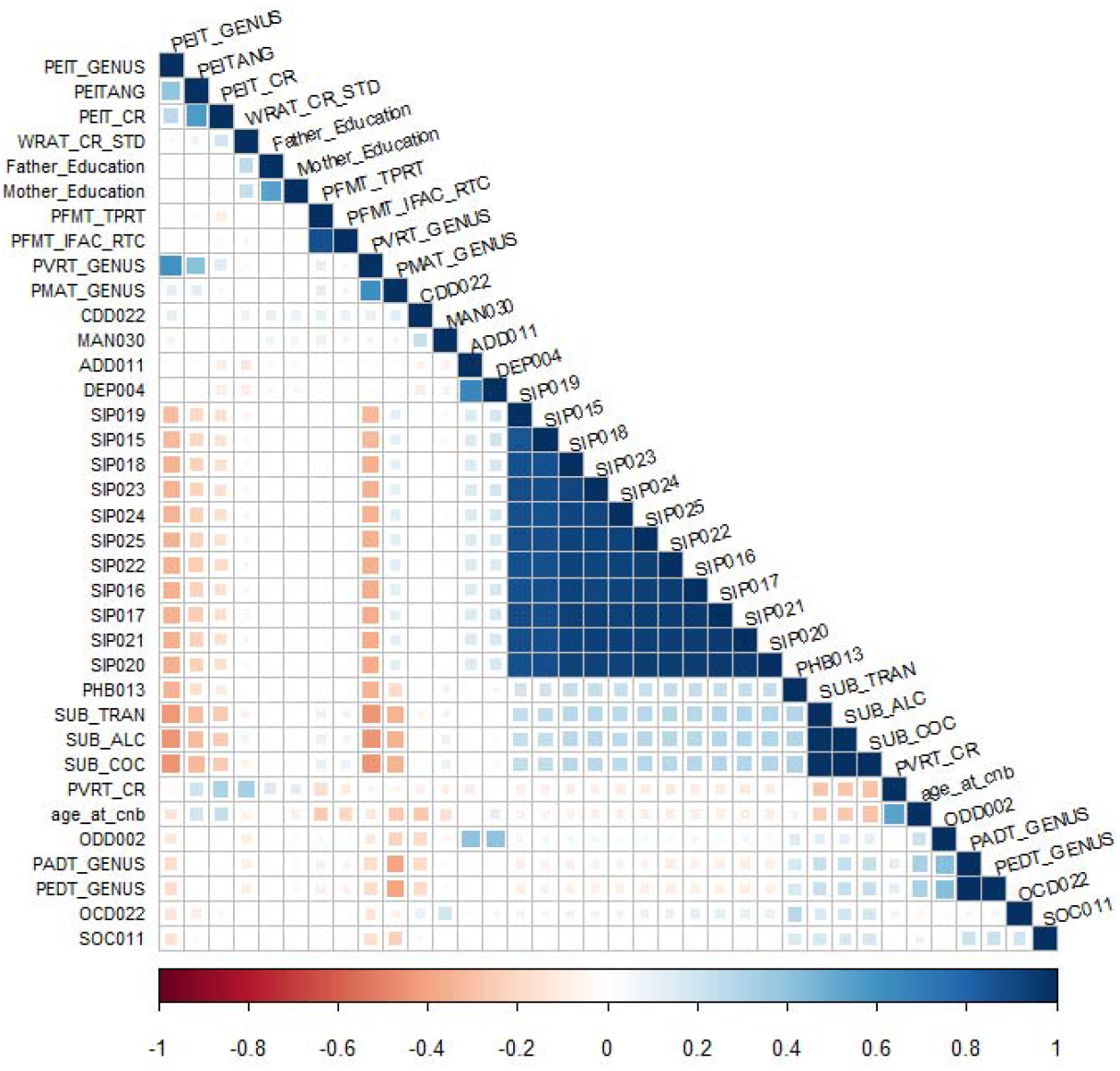
Spearman’s rank-order correlation matrix across 36 neurodevelopmental traits significantly associated with the PRS analyzed. The square size is proportional to the magnitude of the correlation. Blank squares relate to not significant correlations (P>0.01). The legend of the neurodevelopmental codes is reported in Supplemental Table 1.

### Drinks per week

DPW PRS was negatively associated with seven outcomes derived from the SIPS (SIP015: “*I think I have felt that there are odd or unusual things going on that I can’t explain*” Z-score =-3.98, R2=0.4%, P=7.03E-05; SIP016: “*I think that I might be able to predict the future*” Z-score = −3.96, R2=0.4%, P= 7.76E-05; SIP021: “*I wonder if people may be planning to hurt me or even may be about to hurt me*” Z-score = −4.01, R2=0.4%, P= 6.24E-05; SIP022: “*I believe that I have special natural or supernatural gifts beyond my talents and natural strengths*” Z-score = −4.25, R2=0.4%, P= 2.15E-05; SIP023: “*I think I might feel like my mind is “playing tricks” on me*” Z-score = −4.66, R2=0.5%, P= 3.23E-06; SIP024: “*I have had the experience of hearing faint or clear sounds of people or a person mumbling or talking when there is no one near me*” Z-score = −3.91, R2=0.4%, P= 9.27E-05; SIP025: “*I think that I may hear my own thoughts being said out loud*” Z-score = - 3.90, R2=0.4%, P= 9.79E-05). DPW PRS was positively associated with the specific phobia-related item (PHB013: “*Thinking about all of the time that you were afraid of (insert worst fear), whether or not you actually faced it, how long did this fear last? (Weeks)*” Z-score = 3.94, R2=0.4%, P=8.19E-05), one of the obsessive compulsive disorder traits (OCD022 Z-score = 4.00, R2=0.4%, P= 6.42E-05), and PADT_GENUS and PEDT_GENUS (Penn Age Differentiation and Emotion Differentiation Test, both featuring a Z-score = 4.01, R2=0.3%, P= 6.18E-05).

### Age of Smoking Initiation

Age of Smoking Initiation (ASI) PRS was positively associated with the ability to recognize the angry facial emotions of others (PEITANG: *Number of Correct Responses to Anger Trials*, Z-score= 4.61, R2=0.5%, P= 4.14E-06), the standardized score from the Wide Range Achievement Test (Wilkinson et al., 2006) (WRAT_CR_STD: *age-adjusted WRAT test determining participants’ ability to complete the battery and to provide an estimate of IQ*, Z-score = 3.89, R2=0.3%, P= 1.03-04); an item related to conduct disorder traits (CDD022: “*How much did these behaviors change your relationships with your friends?*” Z-score = 3.97, R2=0.3%, P= 7.34-05), and years of father’s education (Z-score = 4.57, R2=0.5%, P= 5.00E-06). The only negative association was found with respect to item SIP019 (“*I think that I may get confused at times whether something I experience or perceive may be real or may be just part of my imagination or dreams*” Z-score = −4.20, R2=0.4%, P= 2.73E-05).

### Smoking Initiation

SI PRS positively associated with the age at completion of the CNB (Z-score = 4.58, R2=0.01%, P=4.70E-06), one of the oppositional defiant disorder traits (ODD002: *“Was there a time when you often got into trouble with adults for refusing to do what they told you to do or for breaking rules at home/school”* Z-score = 4.41, R2=0.7%, P=1.03E-05), a depression-related item (DEP004: “*Has there ever been a time when you felt grouchy, irritable or in a bad mood most of the time; even little things would make you mad?”* Z-score = 4.34, R2=0.6%, P=1.45E-05), and attention deficit disorder (ADD011: *“Did you often have trouble paying attention or keeping your mind on your school, work, chores, or other activities that you were doing?”* Z-score = 4.27, R2=0.6%, P=1.93E-05). A negative association was observed for SI PRS and the years of mother’s education (Z-score =-4.38, R2=0.4%, P=1.22E-05), social anxiety (SOC011: “*Thinking about all of the time that you were afraid of (insert worst fear), whether or not you actually faced it, how long did your fear of this situation last? (Months)”* Z-score = −4.05, R2=0.4%, P=5.21E-05), and a mania-related item (MAN030: *“How much did your feeling (too happy/excited/grouchy/energetic) upset or bother you?”* Z-score = −3.93, R2=0.4%, P=8.48E-05). The two highly correlated phenotypes accounting for the Penn Face Memory Test are also negatively associated with SI PRS (PFMT_TPRT: *Median Response Time for Correct Responses to Target Faces* Z-score = −4.12, R2=0.4%, P=3.86E-05, and PFMT_IFAC_RTC: *Median Response Time for Total Correct Test Trial Responses* Z-score = −4.30, R2=0.4%, P=1.77E-05, respectively). SI PRS was negatively associated with the alcohol (Z-score =-5.31, R2=0.6%, P=1.16E-07), cocaine (Z-score = −5.27, R2=0.6%, P=1.42E-07), and tranquilizer (Z-score = −4.95, R2=0.5%, P=7.85E-07) endorsement phenotypes.

### Cigarettes per Day

CPD PRS was positively associated with three SIPS-derived psychosis outcomes (SIP017: *“I may have felt that there could possibly be something interrupting or controlling my thoughts, feelings, or actions”* Z-score = 4.08, R2=0.4%, P=4.54E-05, SIP018: *“I have had the experience of doing something differently because of my superstitions”* Z-score = 4.04, R2=0.4%, P=5.48E-05, and SIP020: *“I have thought that it might be possible that other people can read my mind, or that I can read others’ minds”* Z-score = 4.00, R2=0.4%, P=6.37E-05. Furthermore, the Penn Emotion Identification Test-related traits and those included in the CNB for the Penn Verbal Reasoning Test were negatively associated to CPD PRS (PEIT_GENUS Z-score = −5.64, R2=0.7%, P=1.87E-08, and PEIT_CR Z-score = −4.37, R2=0.4%, P=1.28E-05, PVRT_GENUS Z-score = −6.11, R2=0.8%, P=1.08E-09, and PVRT_CR Z-score = −4.30, R2=0.3%, P=1.77E-05).

### Smoking Cessation

A unique association was found for SC PRS with the ability to perform the Penn Matrix Reasoning Test, which assesses reasoning by geometric analogy and contrast principle (PMAT_GENUS Z-score = 4.94, R2=0.5%, P=8.06E-07).

### Independence of PRS associations with respect to the genetic overlap among alcohol drinking, tobacco smoking, psychopathology, and other behavioral traits

In order to test whether the PRS associations with neurodevelopmental traits were due to the genetic overlap with psychopathology, educational attainment, cognitive performance, general risk tolerance, and socioeconomic status, their individual PRS were set as additional covariates in the prediction model (Table 1). Building upon the initial regression model (Model-1 covariates: sex, age, the first ten within-ancestry PCs, medical rating, and type of interview), we added the PRS of the other alcohol-drinking and tobacco-smoking traits as covariates (Model-2). In the Model-3, the PRS of psychopathology (i.e., PGC-CD), educational attainment, cognitive performance, general risk tolerance, and socioeconomic status (i.e., HI) were added to the set of covariates included in the Model-2. The significant PRS associations observed in the Model-1 remained significant in the Model-2 and Model-3 analysis with the exception of the association of ASI PRS with WRAT_CR_STD (“*age-adjusted WRAT test determining participants ability to complete the battery and to provide an estimate of IQ*”) when the Model-3 covariates were applied (Model-1: Z-score=3.89, R^2^=0.35%, P=1.03E-04; Model-3: Z-score=1.47, R^2^=0.05%, P=0.142). Remarkably, the associations with substance endorsements and the age at test administration show an increase in magnitude where the adjunctive covariates were considered, suggesting the refinement of the effect when multiple factors were accounted for. To further test the independence of the PRS association from the effects of psychoactive substance use, we removed the participants endorsing the use of alcohol or drugs (N=964; 21%). Applying the Model-1 covariates, we observed consistent PRS associations results between the full PNC cohort and the sample excluding substance users (Figure 4; Supplementary Table 7).

**Figure 4:**
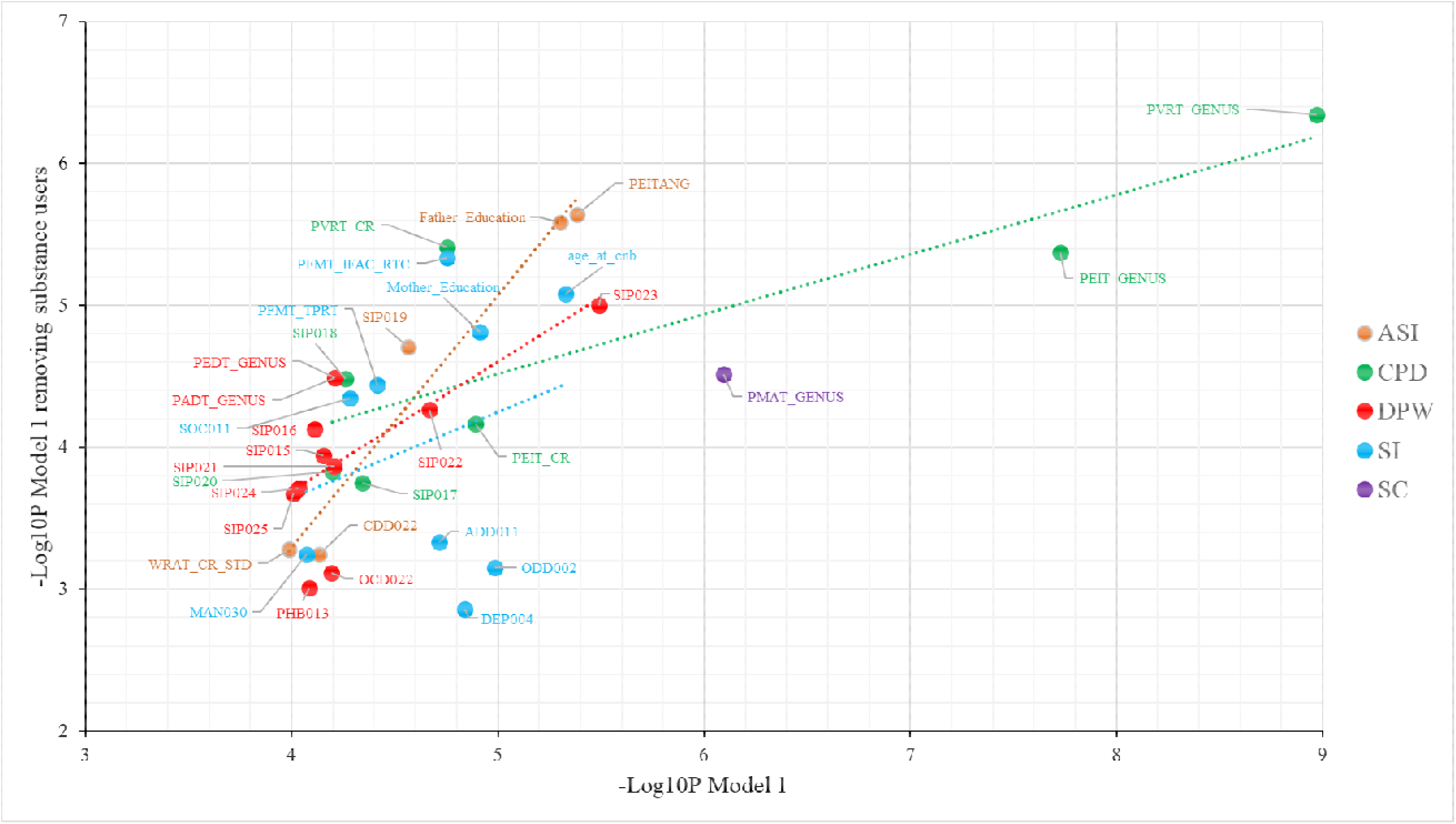
Relationship between the significance of Model 1 and Model 1 excluding substance users associations. The legend of the neurodevelopmental codes is reported in Supplemental Table 1. The dashed lines indicate the linear association between the two models.

### Enrichment Analysis

We interrogated the biological processes enriched related to alcohol-drinking and tobacco-smoking PRS associations, identifying several GO enrichments surviving Bonferroni multiple testing correction (Supplementary Table 8) and applied REVIGO (Supek et al., 2011) to remove redundant terms based on their semantic similarities. The genetic liability for CPD associations involved SNPs enriched for several biological domains, including *neuromuscular junction development* (GO:0007528, P=3.50E-15) and *amyloid precursor protein metabolic process* (GO:0042982, P=1.68E-13), among the most significant. Strong enrichments related to immune systems functions were also observed, including *leukocyte activation involved in inflammatory response* (GO:0002269, P=1.62E-13), *positive regulation of interleukin-6 biosynthetic process* (GO:0045410, P=2E-13), and *erythrocyte maturation* (GO:0043249, P=2.61E-13) (Figure 5). The SNPs underpinning the ASI PRS associations with neurodevelopmental traits were enriched for several biological pathways, whose primary roles seemed to be linked to the muscle tissue and brain development, including *myotube cell development* (GO:0014904, P=4.00E-8), *striated muscle cell differentiation* (GO:0051146, P=5.53E-8), and *response to manganese ion* (GO:0010042, P=7.17E-8). The DPW PRS associations were enriched for terms including *negative regulation of cell projection organization* (GO:0031345, P=5.60E-8) and negative regulation of cell development (GO:0031345, P=6.54E-8). For the SI PRS associations, SNPs were significantly enriched for terms related to diverse functions, including *DNA geometric change* (GO:0032392, P=2.34E-13), *cellular response to carbohydrate stimulus* (GO:0071322, P=5.13E-12), *axo-dendritic protein transport* (GO:0099640, P=1.75E-10), *positive regulation of Phospholipase A2 activity* (GO:0032430, P=2.67E-10), *phosphatidylethanolamine achylchain remodeling* (GO:0036152, P=1.05E-10), and *intracellular receptor signaling pathway* (GO:0030522, P=1.13E-10). Finally, no enrichment related to the SC PRS association survived multiple testing correction.

**Figure 5:**
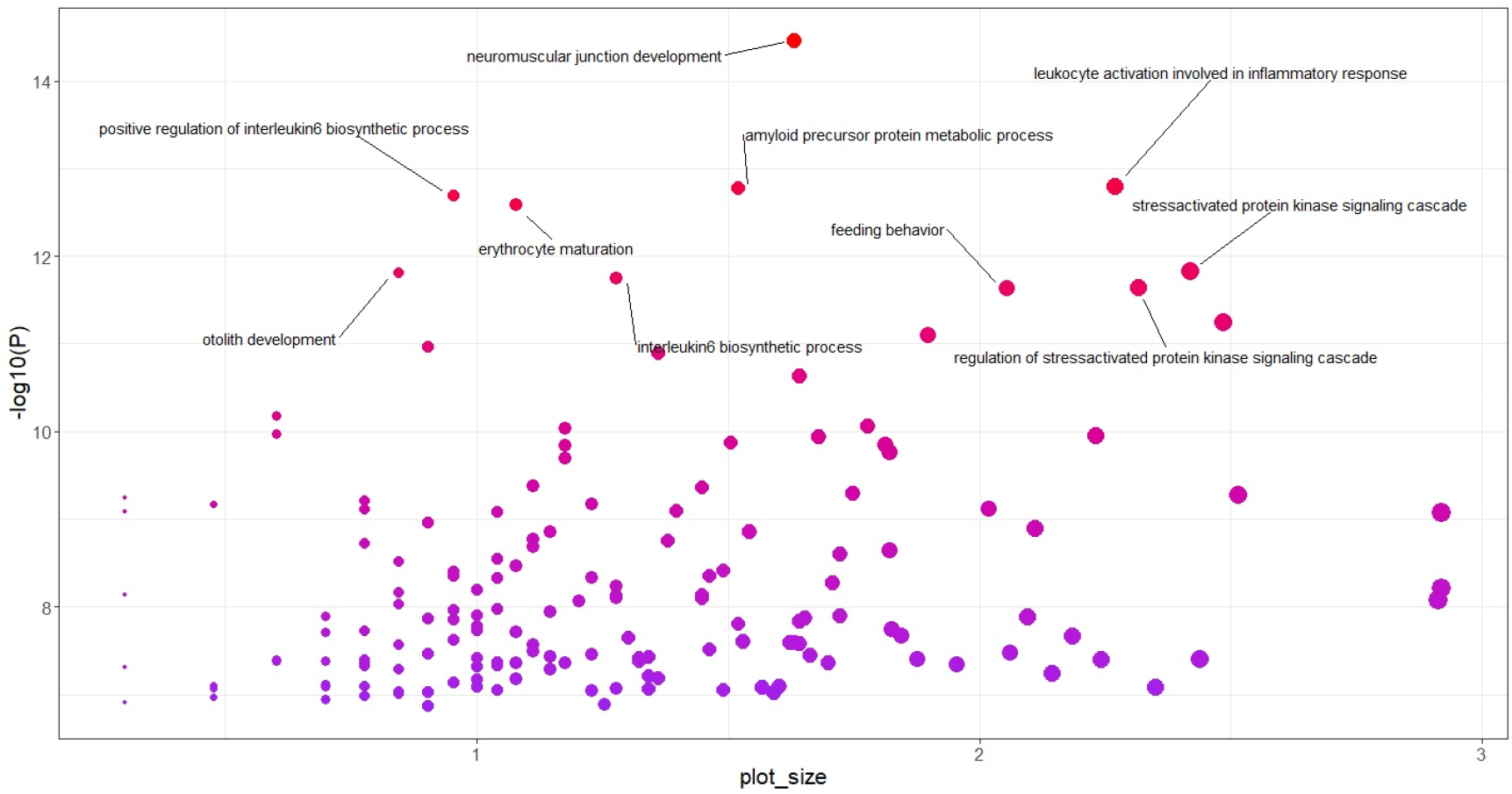
Biological processes enriched in CPD PRS association. Only the processes with more than 10 SNP and surviving the Bonferroni 5% thresholds are labelled. Plot_size indicates the binned frequency of the GO term in the underlying dataset, whit bubbles of more general terms being larger.

## Discussion

Leveraging well-powered genome-wide information generated by the GSCAN study of alcoholic drinks-per-week and four traits related to cigarette consumption (Liu et al., 2019), we investigated the genetic liability for these five traits with respect to neurodevelopmental traits in children and young adults. Furthermore, we tested the independence of the associations observed from potentially correlated genetic effects associated with psychopathology and behavioral traits, as well as effects associated with substance use. We observed a wide range of associations and the majority of them were not affected by the genetic overlap with correlated psychiatric and behavioral traits. Specifically, the association of alcohol-drinking and tobacco-smoking PRS with neurodevelopmental traits was not due to the genetic overlap with psychopathology spectrum, risk tolerance, education, and socioeconomic status, or to substance use among PNC participants. Since the genetic overlap of alcohol-drinking and tobacco-smoking traits with neurodevelopmental traits also appear to be largely mutually independent, we discussed the results of each PRS investigated separately.

### Drinks per Week

This was the only alcohol-related trait analyzed. The DPW PRS was negatively associated with the outcome of several SIPS items capturing the time-length of the specific symptoms as part of an assessment of psychosis presence and severity. The SIPS is a diagnostic interview designed to assess systematically symptoms in individuals who may be at risk for psychosis (Miller et al., 1999; Griffa et al., 2019) that may be prodromal to a psychosis syndrome. Our results indicated that children and young adults with low genetic liability to alcohol drinking reported longer periods of prodromal symptoms. This may appear to diverge from the positive association between alcohol use and psychotic disorders that are supported by hypotheses related to so-called “self-medication” (i.e., individuals with psychotic symptoms use psychoactive substances because they believe they reduce their negative condition) and diathesis-stress model (i.e., genetic predisposition to psychotic disorders interacts with substance use and this interaction results in the onset of schizophrenia) (Polimanti et al., 2017). Alternatively, this may also reflect the genetic differences between quantity-frequency measures of alcohol use, like DPW, and measures of physiological dependence, like alcohol use disorder (Kranzler et al., 2019; Zhou et al., 2020).

As mentioned above, the associations reported were independent of genetic liability to other available traits in the psychopathology spectrum and smoking and drinking habits. Accordingly, we hypothesize that these associations are due to horizontal pleiotropy, i.e., shared molecular pathways between the traits. The main biological processes underlying the DPW associations correspond to the negative regulation of critical cell activities such as the development and the projection organization, ultimately impacting brain activity. Aberrations in these processes have previously been implicated in neurodevelopmental changes underlying psychosis (Leong et al., 2016; Muller et al., 2017; Haarsma et al., 2020) and in altered synapses in the limbic brain areas that drive drinking behavior (Marchant et al., 2016; Sayette, 2017). DPW predisposition was also associated with the number of test trials administered to evaluate the social cognition and neurobehavioral function in Emotion Differentiation and Age Differentiation psychometric tests (Swagerman et al., 2016; Gur et al., 2017). Differences in face recognition are observed in several neurodevelopmental and psychiatric phenotypes (Oruc et al., 2019), and the functional connectivity of the amygdala network is thought to be a key substrate for these functions (Wang et al., 2020). Finally, DPW PRS was also positively associated with anxiety-related obsessive-compulsive disorder. The mesolimbic dopaminergic pathway originating in the ventral tegmental area (VTA) is critical for the onset of reward processing and emotional responses related to anxiety-related disorders (Sun et al., 2019). This evidence could cope with the functional alteration of the VTA projections regulating the alcohol drinking behavior in mice (Juarez et al., 2017), supporting a shared genetic liability.

### Age of Smoking Initiation

Individuals with high genetic liability to early smoking initiation – a trait that is associated clinically with greater severity of affection -- had a longer duration of psychosis symptoms. Despite the well-known association between psychotic illnesses, in particular schizophrenia, and cigarette smoking, a previous study suggested that individuals with psychosis started smoking at a similar age as non-psychotic comparison subjects (Gurillo et al., 2019). Our findings are consistent with the hypothesis that psychosis and the predisposition to early smoking share some genetic liability. ASI PRS is, unsurprisingly, positively associated with the years of paternal education. Adolescents whose parents had less, or no, college education are much more likely to smoke and to smoke earlier than those whose parents have a higher education (Johnston et al., 2016). Our results showed that the association between ASI PRS and father’s education held even when the polygenic components of educational attainment, cognitive performance, and socioeconomic status were added as covariates in the model. This suggests that other mechanisms (e.g., dynastic effects and assortative mating) may be responsible for the genetic overlap between tobacco-smoking behaviors with parents’ education.

ASI PRS was positively associated with higher scores for correct recognition angry faces (PEITANG) when given the options of angry, fearful, happy, neutral, or sad as delivered in the Penn Emotional Identification Test. This relationship suggests that genetic predisposition for smoking initiation may share some liability toward preferential processing of negative social information or perhaps a heightened experience of interpersonal stressors (Brondolo et al., 2003; Kahler et al., 2012). This association was enriched for biological processes mainly related to cellular response to manganese ions (Mn^2+^), which are accumulated in the basal ganglia and are considered a key element in the development of brain activation induced by chronic psychosocial stress in mice (Bouabib et al., 2016; Laine et al., 2017).

The genetic predisposition to later smoking initiation was also positively associated with the ratings of the effects of conduct disorder impacting social relationships (i.e., referencing a pattern of disruptive and violent behaviors following rule-breaking encounters; CDD022: “*How much did these behaviors change your relationships with your friends*?”) suggesting a more consciousness of their disruptive and antisocial behavior in people starting smoking later. A shared genetic etiology among substance abuse and conduct disorder has been previously described (Grant et al., 2015), and our data extend this relationship to the age of smoking initiation.

The Wide Range Assessment Test total standardized score (WRAT_CR_STD) was positively associated with ASI PRS. This test measures individual’s basic skills necessary for effective learning, communication, and thinking, such as reading words, comprehending sentences, spelling out, and solving math problems (Harvey et al., 2006), and was used to triage individuals for their anticipated ability to complete the PNC testing battery. An impact of smoking on cognitive decline has been observed in adulthood (Sabia et al., 2008; Sabia et al., 2012) and childhood (Julvez et al., 2007). Nonetheless, our study suggests that this relationship might be also due to a shared genetic predisposition rather than the sole effect of tobacco smoking. However, this association is no longer significant when the covariate PRS of cognitive performance, education attainment, household income, risk-taking behavior, are included in the model along with the psychopathology liability. This suggests that the genetic overlap between ASI and WRAT_CR_STD is driven by another genetically correlated trait.

While early tobacco use was already associated with specific non-affective psychosis (McGrath et al., 2016), the association of ASI PRS with SIP019 phenotype (i.e., “*I think that I may get confused at times whether something I experience or perceive may be real or maybe just part of my imagination or dreams*”) supports a partial contribution from horizontal pleiotropy (i.e., shared genetic basis) between these traits.

### Cigarettes per Day

CPD PRS was positively associated with several SIPS-items indicating the duration of prodromal psychotic symptoms. This finding is consistent with the shared genetic liability observed in discordant twin and sibling studies of schizophrenia (Kendler et al., 2015), where smoking prospectively predicts risk for schizophrenia. The SNPs involved in the present associations were enriched with multiple biological processes related to cellular signaling homeostasis, including terms related to *Synaptic Vesicles Membrane* (GO:0030672) and *Signal Release* (GO:0023061), that might impact the neuronal activity. The synaptic plasticity in brain areas modulates the activity of neural circuitries, and the disruption of this activity might result in detrimental consequences underlying psychotic behaviors (Crabtree et al., 2014; Mizuno et al., 2019).

The genetic liability to smoking quantity was negatively associated with emotional identification and verbal reasoning independently from psychopathology, substance use, and other behavioral traits. Accordingly, we can hypothesize that certain molecular mechanisms involved in the predisposition to smoking quantity are shared with these neurodevelopmental traits.

We observed multiple neuroinflammatory pathways among the biological processes enriched for SNPs mediating the CPD PRS associations. Interleukin-6 is a pro-inflammatory cytokine with wide-ranging biological effects. It has been previously implicated in neuroinflammation in the context of neuropsychiatric disorders and also appears to correlate with neurocognitive changes in the context of aging (Bradburn et al., 2017; Fominykh et al., 2018; Bobbo et al., 2019; Jung et al., 2019). Experimental manipulations of IL-6 signaling appear capable of producing related effects in animal models (Donegan et al., 2014, Hodes et al., 2014). While acute tobacco smoke exposure was previously associated with attenuated IL-6 signaling in alveolar macrophages, chronic smoking appears to be associated with higher circulating IL-6 in a large multi-site observational study, in which an interaction between smoking history and a SNP in the IL-6 promotor was predictive of circulating IL-6 and CRP levels (Sunyer et al., 2009). Thus, it seems plausible that pleiotropic effects on smoking quantity and aspects of cognitive development may involve the IL-6 pathway. However, controlling for genetic liabilities for cognitive performance, psychopathology, and removing effects associated with reported exposure to substances of abuse did not alter this association. Moreover, the genetic liability to smoking more cigarettes per day was previously shown to be enriched with biological processes related to amyloid metabolism and neuroinflammation (Winer et al., 2018).

### Smoking Initiation and Cessation

Genetic predisposition to smoking initiation was positively associated with items related to oppositional defiant disorder in children and young adults. The positive phenotypic correlation between ODD002 (Oppositional Defiant Disorder: *Was there a time when you often got into trouble with adults for refusing to do what they told you to do or for breaking rules at home/school?*) and both ADD011 (Attention Deficit Disorder: *Did you often have trouble paying attention or keeping your mind on your school, work, chores, or other activities that you were doing?*) and DEP004 (Depression: *Has there ever been a time when you felt grouchy, irritable or in a bad mood most of the time; even little things would make you mad?*) were already described (Noordermeer et al., 2017; Ding et al., 2020) and might account for the associations’ consistency with SI PRS. It is also consistent with the fact that initiating smoking is, itself, often and oppositional/defiant behavior. The dysregulations of dopaminergic and nicotinic pathways are shared mechanisms between smoking habits and the onset of attention deficit disorders (van Amsterdam et al., 2018). Conversely, the relationship between tobacco smoking and depressive status and anxiety is inconsistent in terms of the direction of association (Fluharty et al., 2017), suggesting the need for future explanations. Our results supported the sharing of genetic determinants for smoking initiation and these survey item reports, even when the genetic predisposition for psychopathology was included as a covariate in the model. Nevertheless, environmental factors, such as the family context and parental behaviors, play a crucial role in the onset of these behaviors (Deault, 2010; Banerjee et al., 2011; Thapar et al., 2013). Indeed, higher levels of maternal education were associated with lower likelihood of being a smoker (Moore et al., 2015). This association held on even after we included PRSs for psychosocial and psychopathological traits in the model, supporting the role of the familiar factors in primarily influence the SI genetic predisposition in early adolescence (Maes et al., 2017).

Two more neurodevelopmental traits, SOC011 (Social Anxiety: *Thinking about all of the time that you were afraid of your worst fear, whether or not you actually faced it, how long did your fear of this situation last in months?)* and the age when the computerized neurocognitive battery was completed, were associated with SI PRS, and, as expected, the SI PRS is also positively related to the substance use. A child with a higher genetic vulnerability for smoking initiation may have more behavioral issues in general (Minichino et al., 2013), which might prompt parents to seek evaluation at an earlier age. The positive relationship between the SI PRS and age at neurocognitive testing was enriched with genetic variants involved in brain-relevant biological pathways, including positive regulation of the phospholipase A2 (PLA2) (GO: 0032430), which is involved in a pro-inflammatory status via the release of arachidonic acid, acting pivotally in the intracellular membrane trafficking and apoptotic processes (Sun et al., 2004). It may also contribute to nervous system degeneration (Sun et al., 2004; Collins et al., 2014), and appears to impact iron accumulation in the globus pallidus, the substantia nigra and the dentate nucleus (Lehéricy et al., 2020). Our data are consistent with the previously reported association between PLA2 genetic variation and the development of smoking habits in people affected by psychiatric disorders (Nadalin et al., 2019). Other genetic variants underpinning this association are also involved in the axodendritic protein transport (GO: 0099640), suggesting their role in the proper synapse development and prompting the phospholipase A2 activity as relevant for brain development and synaptic functioning (Law et al., 2006).

Our data confirm that tobacco smoking and alcohol drinking share a genetic liability involving multiple biological processes (Prom-Wormley et al., 2017), and the most significant functional enriched process in substance use association was the DNA geometric change (GO:0032392). Genetic variants underlying substance use associations recognized cellular response to carbohydrate stimuli, that are increasingly considered to alter brain circuitry, leveraging the induction of dopamine reward and craving that are comparable in magnitude to those induced by addictive drugs or alcohol use (Gordon et al., 2018;Winterdahl et al., 2019).

The substance associations with SI PRS were also driven by genetic variants involved in the metabolism of phosphatidylethanolamine, a phospholipid critical for white matter establishment (Fledrich et al., 2018; Unterrainer et al., 2019).

Finally, SC PRS shares the genetic liability with the Penn Matrix Reasoning Test. This test measures nonverbal reasoning ability assessed via the completion of matrices requiring reasoning by geometric analogy and contrast principles (Moore et al., 2015). Our data suggest that people who opted for fewer practice trials before the test showed less genetic predisposed to quit smoking.

## Conclusions

Our study provided evidence that the polygenic risk for smoking and alcohol-use phenotypes overlap with several neurodevelopmental traits assessed in a population-based cohort of children and adolescence, and that these relationships appeared independent of actual psychoactive substance use. The associations were also independent from the genetic effects exerted by genetically correlated phenotypes, including other substance use phenotypes, psychopathology and psychosocial factors. Our findings highlight plausible pleiotropic mechanisms linking genetic liability to smoking and drinking behaviors to aspects of cognitive and behavioral development.

## Data Availability

All the data are available in the manuscript

## Abbreviations

DPW: Drinks Per Week
ASI: Age of Smoking Initiation
SI: Smoking Initiation
CPD: Cigarettes Per Day
SC: Smoking Cessation
PGC-CD: Psychiatric Genomics Consortium Cross Disorder
CP: Cognitive Performance
EA: Educational Attainment
GR: General Risk-taking behavior
HI: Household Income
PNC: Philadelphia Neurodevelopmental Cohort

## Key points and relevance

- The polygenic risk for smoking and alcohol-use phenotypes overlap with several neurodevelopmental traits assessed in a population-based cohort of children and adolescence
- These relationships appeared independent of actual psychoactive substance use.
- The associations were independent from the genetic effects exerted by other substance use phenotypes, psychopathology and psychosocial factors.
- Our findings highlight plausible pleiotropic mechanisms linking genetic liability to these hazardous behaviors to aspects of cognitive and behavioral development, representing a relevant contribution to the topic enhancing further science investigation.

## Acknowledgements

We would like to acknowledge support from the National Institutes of Health (R21 DA047527, R21 DC018098, F32 MH122058).

## Notes

### Competing Interest Statement

The authors have declared no competing interest.

### Funding Statement

We would like to acknowledge support from the National Institutes of Health (R21 DA047527, R21 DC018098, F32 MH122058) allotted to RP.

### Author Declarations

We use anonymized data, so no IRB approval is necessary.

## References

Alzola, C., & Harrell, F. (s.d.). An Introduction to S and the Hmisc and Design Libraries. 310.

Banerjee, J., Bhojani, S., & Emcy, N. (2011). Co-existence of ADHD, autoimmune hypothyroidism and pituitary macroadenoma presenting in a behaviour clinic: A case report and brief review of the literature. Journal of Pediatric Endocrinology and Metabolism, 24(3–4), 229–231. https://doi.org/10.1515/jpem.2011.122

Bobbo, V. C. D., Jara, C. P., Mendes, N. F., Morari, J., Velloso, L. A., & Araújo, E. P. (2019). Interleukin-6 Expression by Hypothalamic Microglia in Multiple Inflammatory Contexts: A Systematic Review. BioMed Research International, 2019, 1365210. https://doi.org/10.1155/2019/1365210

Bouabid, S., Tinakoua, A., Lakhdar[Ghazal, N., & Benazzouz, A. (2016). Manganese neurotoxicity: Behavioral disorders associated with dysfunctions in the basal ganglia and neurochemical transmission. Journal of Neurochemistry, 136(4), 677–691. https://doi.org/10.1111/jnc.13442

Brondolo, E., Rieppi, R., Erickson, S. A., Bagiella, E., Shapiro, P. A., McKinley, P., & Sloan, R. P. (2003). Hostility, Interpersonal Interactions, and Ambulatory Blood Pressure. Psychosomatic Medicine, 65(6), 1003–1011. https://doi.org/10.1097/01.PSY.0000097329.53585.A1

Bulik-Sullivan, B., Finucane, H. K., Anttila, V., Gusev, A., Day, F. R., Loh, P.-R., Duncan, L., Perry, J. R. B., Patterson, N., Robinson, E. B., Daly, M. J., Price, A. L., & Neale, B. M. (2015). An atlas of genetic correlations across human diseases and traits. Nature Genetics, 47(11), 1236–1241. https://doi.org/10.1038/ng.3406

Castanon, N., Luheshi, G., & Layé, S. (2015). Role of neuroinflammation in the emotional and cognitive alterations displayed by animal models of obesity. Frontiers in Neuroscience, 9, 229. https://doi.org/10.3389/fnins.2015.00229

Chang, C. C., Chow, C. C., Tellier, L. C., Vattikuti, S., Purcell, S. M., & Lee, J. J. (2015). Second-generation PLINK: Rising to the challenge of larger and richer datasets. GigaScience, 4(1). https://doi.org/10.1186/s13742-015-0047-8

Choi, S. W., & O’Reilly, P. F. (2019). PRSice-2: Polygenic Risk Score software for biobank-scale data. GigaScience, 8(7). https://doi.org/10.1093/gigascience/giz082

Clarke, T.-K., Adams, M. J., Davies, G., Howard, D. M., Hall, L. S., Padmanabhan, S., Murray, A. D., Smith, B. H., Campbell, A., Hayward, C., Porteous, D. J., Deary, I. J., & McIntosh, A. M. (2017). Genome-wide association study of alcohol consumption and genetic overlap with other health-related traits in UK Biobank (N =112 117). Molecular Psychiatry, 22(10), 1376–384. https://doi.org/10.1038/mp.2017.153

Collins, M. A., Tajuddin, N., Moon, K.-H., Kim, H.-Y., Nixon, K., & Neafsey, E. J. (2014). Alcohol, phospholipase A2-associated neuroinflammation, and ω3 docosahexaenoic acid protection. Molecular Neurobiology, 50(1), 239–245. https://doi.org/10.1007/s12035-014-8690-0

Couto, F. M., Silva, M. J., & Coutinho, P. M. (2007). Measuring semantic similarity between Gene Ontology terms. Data & Knowledge Engineering, 61(1), 137–152. https://doi.org/10.1016/j.datak.2006.05.003

Crabtree, G. W., & Gogos, J. A. (2014). Synaptic plasticity, neural circuits, and the emerging role of altered short-term information processing in schizophrenia. Frontiers in Synaptic Neuroscience, 6. https://doi.org/10.3389/fnsyn.2014.00028

Cross-Disorder Group of the Psychiatric Genomics Consortium, Lee, P. H., Anttila, V., Won, H., Feng, Y.-C. A., Rosenthal, J., Zhu, Z., Tucker-Drob, E. M., Nivard, M. G., Grotzinger, A. D., Posthuma, D., Wang, M. M.-J., Yu, D., Stahl, E., Walters, R. K., Anney, R. J. L., Duncan, L. E., Belangero, S., Luykx, J., … Smoller, J. W. (2019). Genome wide meta-analysis identifies genomic relationships, novel loci, and pleiotropic mechanisms across eight psychiatric disorders. BioRxiv, 528117. https://doi.org/10.1101/528117

Deault, L. C. (2010). A systematic review of parenting in relation to the development of comorbidities and functional impairments in children with attention-deficit/hyperactivity disorder (ADHD). Child Psychiatry and Human Development, 41(2), 168–192. https://doi.org/10.1007/s10578-009-0159-4

Ding, W., Meza, J., Lin, X., He, T., Chen, H., Wang, Y., & Qin, S. (2020). Oppositional Defiant Disorder Symptoms and Children’s Feelings of Happiness and Depression: Mediating Roles of Interpersonal Relationships. Child Indicators Research, 13(1), 215–235. https://doi.org/10.1007/s12187-019-09685-9

Erzurumluoglu, A. M., Liu, M., Jackson, V. E., Barnes, D. R., Datta, G., Melbourne, C. A., Young, R., Batini, C., Surendran, P., Jiang, T., Adnan, S. D., Afaq, S., Agrawal, A., Altmaier, E., Antoniou, A. C., Asselbergs, F. W., Baumbach, C., Bierut, L., Bertelsen, S., … Howson, J. M. M. (2019). Meta-analysis of up to 622,409 individuals identifies 40 novel smoking behaviour associated genetic loci. Molecular Psychiatry, 1–18. https://doi.org/10.1038/s41380-018-0313-0

Finucane, H. K., Bulik-Sullivan, B., Gusev, A., Trynka, G., Reshef, Y., Loh, P.-R., Anttila, V., Xu, H., Zang, C., Farh, K., Ripke, S., Day, F. R., Purcell, S., Stahl, E., Lindstrom, S., Perry, J. R. B., Okada, Y., Raychaudhuri, S., Daly, M. J., … Price, A. L. (2015). Partitioning heritability by functional annotation using genome-wide association summary statistics. Nature Genetics, 47(11), 1228–1235. https://doi.org/10.1038/ng.3404

Fledrich, R., Abdelaal, T., Rasch, L. et al. Targeting myelin lipid metabolism as a potential therapeutic strategy in a model of CMT1A neuropathy. Nat Commun 9, 3025 (2018). https://doi.org/10.1038/s41467-018-05420-0

Fluharty, M., Taylor, A. E., Grabski, M., & Munafò, M. R. (2017). The Association of Cigarette Smoking With Depression and Anxiety: A Systematic Review. Nicotine & Tobacco Research: Official Journal of the Society for Research on Nicotine and Tobacco, 19(1), 3–13. https://doi.org/10.1093/ntr/ntw140

Fominykh, V., Vorobyeva, A., Onufriev, M. V., Brylev, L., Zakharova, M. N., & Gulyaeva, N. V. (2018). Interleukin-6, S-Nitrosothiols, and Neurodegeneration in Different Central Nervous System Demyelinating Disorders: Is There a Relationship? Journal of Clinical Neurology (Seoul, Korea), 14(3), 327–332. https://doi.org/10.3988/jcn.2018.14.3.327

Gelernter, J., Zhou, H., Nuñez, Y. Z., Mutirangura, A., Malison, R. T., & Kalayasiri, R. (2018). Genomewide Association Study of Alcohol Dependence and Related Traits in a Thai Population. Alcoholism, Clinical and Experimental Research, 42(5), 861–868. https://doi.org/10.1111/acer.13614

Gordon, E. L., Ariel-Donges, A. H., Bauman, V., & Merlo, L. J. (2018). What Is the Evidence for «Food Addiction?» A Systematic Review. Nutrients, 10(4). https://doi.org/10.3390/nu10040477

Grant, J. D., Lynskey, M. T., Madden, P. A. F., Nelson, E. C., Few, L. R., Bucholz, K. K., Statham, D. J., Martin, N. G., Heath, A. C., & Agrawal, A. (2015). The role of conduct disorder in the relationship between alcohol, nicotine and cannabis use disorders. Psychological medicine, 45(16), 3505–3515. https://doi.org/10.1017/S0033291715001518

Griffa, A., Baumann, P. S., Klauser, P., Mullier, E., Cleusix, M., Jenni, R., van den Heuvel, M. P., Do, K. Q., Conus, P., & Hagmann, P. (2019). Brain connectivity alterations in early psychosis: From clinical to neuroimaging staging. Translational Psychiatry, 9(1), 1–10. https://doi.org/10.1038/s41398-019-0392-y

Gur, R. E., Moore, T. M., Calkins, M. E., Ruparel, K., & Gur, R. C. (2017). Face Processing Measures of Social Cognition: A Dimensional Approach to Developmental Psychopathology. Biological Psychiatry: Cognitive Neuroscience and Neuroimaging, 2(6), 502–509. https://doi.org/10.1016/j.bpsc.2017.03.010

Gurillo, P., Jauhar, S., Murray, R. M., & MacCabe, J. H. (2015). Does tobacco use cause psychosis? Systematic review and meta-analysis. The Lancet Psychiatry, 2(8), 718–725. https://doi.org/10.1016/S2215-0366(15)00152-2

Haarsma, J., Fletcher, P. C., Griffin, J. D., Taverne, H. J., Ziauddeen, H., Spencer, T. J., Miller, C., Katthagen, T., Goodyer, I., Diederen, K. M. J., & Murray, G. K. (2020). Precision weighting of cortical unsigned prediction error signals benefits learning, is mediated by dopamine, and is impaired in psychosis. Molecular Psychiatry, 1–14. https://doi.org/10.1038/s41380-020-0803-8

Harvey, P. D., Friedman, J. I., Bowie, C., Reichenberg, A., McGurk, S. R., Parrella, M., White, L., & Davis, K. L. (2006). Validity and stability of performance-based estimates of premorbid educational functioning in older patients with schizophrenia. Journal of Clinical and Experimental Neuropsychology, 28(2), 178–192. https://doi.org/10.1080/13803390500360349

Hill, W. D., Davies, N. M., Ritchie, S. J., Skene, N. G., Bryois, J., Bell, S., Di Angelantonio, E., Roberts, D. J., Xueyi, S., Davies, G., Liewald, D. C. M., Porteous, D. J., Hayward, C., Butterworth, A. S., McIntosh, A. M., Gale, C. R., & Deary, I. J. (2019). Genome-wide analysis identifies molecular systems and 149 genetic loci associated with income. Nature Communications, 10(1), 5741. https://doi.org/10.1038/s41467-019-13585-5

Howie, B. N., Donnelly, P., & Marchini, J. (2009). A Flexible and Accurate Genotype Imputation Method for the Next Generation of Genome-Wide Association Studies. PLOS Genetics, 5(6), e1000529. https://doi.org/10.1371/journal.pgen.1000529

Johnston LD, O’Malley PM, Miech RA, Bachman JG, Schulenberg JE. Monitoring The Future National Survey Results On Drug Use, 1975-2015: Overview Of Key Findings On Adolescent Drug Use. Institute for Social Research, The University of Michigan; Ann Arbor, MI: 2016. Available at http://monitoringthefuture.org/pubs/monographs/mtf-overview2015.pdf.

Jorgenson, E., Thai, K. K., Hoffmann, T. J., Sakoda, L. C., Kvale, M. N., Banda, Y., Schaefer, C., Risch, N., Mertens, J., Weisner, C., & Choquet, H. (2017). Genetic contributors to variation in alcohol consumption vary by race/ethnicity in a large multi-ethnic genome-wide association study. Molecular Psychiatry, 22(9), 1359–1367. https://doi.org/10.1038/mp.2017.101

Juarez, B., Morel, C., Ku, S. M., Liu, Y., Zhang, H., Montgomery, S., Gregoire, H., Ribeiro, E., Crumiller, M., Roman-Ortiz, C., Walsh, J. J., Jackson, K., Croote, D. E., Zhu, Y., Zhang, S., Vendruscolo, L. F., Edwards, S., Roberts, A., Hodes, G. E., … Han, M.-H. (2017). Midbrain circuit regulation of individual alcohol drinking behaviors in mice. Nature Communications, 8(1), 2220. https://doi.org/10.1038/s41467-017-02365-8

Julvez, J., Ribas-Fitó, N., Torrent, M., Forns, M., Garcia-Esteban, R., & Sunyer, J. (2007). Maternal smoking habits and cognitive development of children at age 4 years in a population-based birth cohort. International Journal of Epidemiology, 36(4), 825–832. https://doi.org/10.1093/ije/dym107

Jung, Y.-H., Shin, N. Y., Jang, J. H., Lee, W. J., Lee, D., Choi, Y., Choi, S.-H., & Kang, D.-H. (2019). Relationships among stress, emotional intelligence, cognitive intelligence, and cytokines. Medicine, 98(18). https://doi.org/10.1097/MD.0000000000015345

Kahler, C. W., McHugh, R. K., Leventhal, A. M., Colby, S. M., Gwaltney, C. J., & Monti, P. M. (2012). High Hostility Among Smokers Predicts Slower Recognition of Positive Facial Emotion. Personality and individual differences, 52(3), 444–448. https://doi.org/10.1016/j.paid.2011.11.009

Karlsson Linnér, R., Biroli, P., Kong, E., Meddens, S. F. W., Wedow, R., Fontana, M. A., Lebreton, M., Tino, S. P., Abdellaoui, A., Hammerschlag, A. R., Nivard, M. G., Okbay, A., Rietveld, C. A., Timshel, P. N., Trzaskowski, M., … Beauchamp, J. P. (2019). Genome-wide association analyses of risk tolerance and risky behaviors in over 1 million individuals identify hundreds of loci and shared genetic influences. Nature Genetics, 51(2), 245–257. https://doi.org/10.1038/s41588-018-0309-3

Kendler, K. S., Lönn, S. L., Sundquist, J., & Sundquist, K. (2015). Smoking and schizophrenia in population cohorts of Swedish women and men: A prospective co-relative control study. The American Journal of Psychiatry, 172(11), 1092–1100. https://doi.org/10.1176/appi.ajp.2015.15010126

Kranzler, H. R., Zhou, H., Kember, R. L., Vickers Smith, R., Justice, A. C., Damrauer, S., Tsao, P. S., Klarin, D., Baras, A., Reid, J., Overton, J., Rader, D. J., Cheng, Z., Tate, J. P., Becker, W. C., Concato, J., Xu, K., Polimanti, R., Zhao, H., & Gelernter, J. (2019). Genome-wide association study of alcohol consumption and use disorder in 274,424 individuals from multiple populations. Nature Communications, 10(1), 1499. https://doi.org/10.1038/s41467-019-09480-8

Laine, M. A., Sokolowska, E., Dudek, M., Callan, S.-A., Hyytiä, P., & Hovatta, I. (2017). Brain activation induced by chronic psychosocial stress in mice. Scientific Reports, 7(1), 15061. https://doi.org/10.1038/s41598-017-15422-5

Law, M. H., Cotton, R. G. H., & Berger, G. E. (2006). The role of phospholipases A2 in schizophrenia. Molecular Psychiatry, 11(6), 547–556. https://doi.org/10.1038/sj.mp.4001819

Lee, J. J., Wedow, R., Okbay, A., Kong, E., Maghzian, O., Zacher, M., Nguyen-Viet, T. A., Bowers, P., Sidorenko, J., Linnér, R. K., Fontana, M. A., Kundu, T., Lee, C., Li, H., Li, R., Royer, R., Timshel, P. N., Walters, R. K., Willoughby, E. A., … Cesarini, D. (2018). Gene discovery and polygenic prediction from a genome-wide association study of educational attainment in 1.1 million individuals. Nature Genetics, 50(8), 1112–1121. https://doi.org/10.1038/s41588-018-0147-3

Lehéricy, S., Roze, E., Goizet, C., & Mochel, F. (2020). MRI of neurodegeneration with brain iron accumulation. Current Opinion in Neurology, 33(4), 462–473. https://doi.org/10.1097/WCO.0000000000000844

Leong, A. T. L., Chan, R. W., Gao, P. P., Chan, Y.-S., Tsia, K. K., Yung, W.-H., & Wu, E. X. (2016). Long-range projections coordinate distributed brain-wide neural activity with a specific spatiotemporal profile. Proceedings of the National Academy of Sciences, 113(51), E8306–E8315. https://doi.org/10.1073/pnas.1616361113

Liu, M., Jiang, Y., Wedow, R., Li, Y., Brazel, D. M., Chen, F., Datta, G., Davila-Velderrain, J., McGuire, D., Tian, C., Zhan, X., Choquet, H., Docherty, A. R., Faul, J. D., Foerster, J. R., Fritsche, L. G., Gabrielsen, M. E., Gordon, S. D., Haessler, J., … Vrieze, S. (2019). Association studies of up to 1.2 million individuals yield new insights into the genetic etiology of tobacco and alcohol use. Nature Genetics, 51(2), 237–244. https://doi.org/10.1038/s41588-018-0307-5

Maes, H. H., Prom-Wormley, E., Eaves, L. J., Rhee, S. H., Hewitt, J. K., Young, S., Corley, R., McGue, M., Iacono, W. G., Legrand, L., Samek, D. R., Murrelle, E. L., Silberg, J. L., Miles, D. R., Schieken, R. M., Beunen, G. P., Thomis, M., Rose, R. J., Dick, D. M., Boomsma, D. I., … Neale, M. C. (2017). A Genetic Epidemiological Mega Analysis of Smoking Initiation in Adolescents. Nicotine & tobacco research : official journal of the Society for Research on Nicotine and Tobacco, 19(4), 401–409. https://doi.org/10.1093/ntr/ntw294

Marchant, N. J., Campbell, E. J., Whitaker, L. R., Harvey, B. K., Kaganovsky, K., Adhikary, S., Hope, B. T., Heins, R. C., Prisinzano, T. E., Vardy, E., Bonci, A., Bossert, J. M., & Shaham, Y. (2016). Role of Ventral Subiculum in Context-Induced Relapse to Alcohol Seeking after Punishment-Imposed Abstinence. Journal of Neuroscience, 36(11), 3281–3294. https://doi.org/10.1523/JNEUROSCI.4299-15.2016

Martin, A. R., Gignoux, C. R., Walters, R. K., Wojcik, G. L., Neale, B. M., Gravel, S., Daly, M. J., Bustamante, C. D., & Kenny, E. E. (2017). Human Demographic History Impacts Genetic Risk Prediction across Diverse Populations. American Journal of Human Genetics, 100(4), 635–649. https://doi.org/10.1016/j.ajhg.2017.03.004

Matoba, N., Akiyama, M., Ishigaki, K., Kanai, M., Takahashi, A., Momozawa, Y., Ikegawa, S., Ikeda, M., Iwata, N., Hirata, M., Matsuda, K., Kubo, M., Okada, Y., & Kamatani, Y. (2019a). GWAS of smoking behaviour in 165,436 Japanese people reveals seven new loci and shared genetic architecture. Nature Human Behaviour, 3(5), 471–477. https://doi.org/10.1038/s41562-019-0557-y

McGrath, J. J., Alati, R., Clavarino, A., Williams, G. M., Bor, W., Najman, J. M., Connell, M., & Scott, J. G. (2016). Age at first tobacco use and risk of subsequent psychosis-related outcomes: A birth cohort study. The Australian and New Zealand Journal of Psychiatry, 50(6), 577–583. https://doi.org/10.1177/0004867415587341

Miller, Tandy J., McGlashan, T. H., Woods, S. W., Stein, K., Driesen, N., Corcoran, C. M., Hoffman, R., & Davidson, L. (1999). Symptom Assessment in Schizophrenic Prodromal States. Psychiatric Quarterly, 70(4), 273–287. https://doi.org/10.1023/A:1022034115078

Minichino, A., Bersani, F. S., Calò, W. K., Spagnoli, F., Francesconi, M., Vicinanza, R., Delle Chiaie, R., & Biondi, M. (2013). Smoking Behaviour and Mental Health Disorders—Mutual Influences and Implications for Therapy. International Journal of Environmental Research and Public Health, 10(10), 4790–4811. https://doi.org/10.3390/ijerph10104790

Mizuno, T., Matsumoto, H., Mita, K., Kogauchi, S., Kiyono, Y., Kosaka, H., & Omata, N. (2019). Psychosis is an extension of mood swings from the perspective of neuronal plasticity impairments. Medical Hypotheses, 124, 37–39. https://doi.org/10.1016/j.mehy.2019.02.001

Moore, T. M., Reise, S. P., Gur, R. E., Hakonarson, H., & Gur, R. C. (2015). Psychometric properties of the Penn Computerized Neurocognitive Battery. Neuropsychology, 29(2), 235–246. https://doi.org/10.1037/neu0000093

Moore, T. M., Reise, S. P., Gur, R. E., Hakonarson, H., & Gur, R. C. (2015a). Psychometric Properties of the Penn Computerized Neurocognitive Battery. Neuropsychology, 29(2), 235–246. https://doi.org/10.1037/neu0000093

Müller, C. P., Kalinichenko, L. S., Tiesel, J., Witt, M., Stöckl, T., Sprenger, E., Fuchser, J., Beckmann, J., Praetner, M., Huber, S. E., Amato, D., Mühle, C., Büttner, C., Ekici, A. B., Smaga, I., Pomierny-Chamiolo, L., Pomierny, B., Filip, M., Eulenburg, V., … Kornhuber, J. (2017). Paradoxical antidepressant effects of alcohol are related to acid sphingomyelinase and its control of sphingolipid homeostasis. Acta Neuropathologica, 133(3), 463–483. https://doi.org/10.1007/s00401-016-1658-6

Nadalin, S., Rebić, J., Šendula Jengić, V., Peitl, V., Karlović, D., & Buretić-Tomljanović, A. (2019b). Association between PLA2G6 gene polymorphism for calcium-independent phospholipase A2 and nicotine dependence among males with schizophrenia. Prostaglandins, Leukotrienes and Essential Fatty Acids, 148, 9–15. https://doi.org/10.1016/j.plefa.2019.07.005

Nana, M., Link to external site, this link will open in a new window, Masato, A., Kazuyoshi, I., Masahiro, K., Link to external site, this link will open in a new window, Takahashi, A., Yukihide, M., Shiro, I., Masashi, I., Nakao, I., Makoto, H, Koichi, M., Michiaki, K., Yukinori, O., … (2019). GWAS of smoking behaviour in 165,436 Japanese people reveals seven new loci and shared genetic architecture. Nature Human Behaviour; London, 3(5), 471–477. http://dx.doi.org/10.1038/s41562-019-0557-y

Noordermeer, S. D. S., Luman, M., Weeda, W. D., Buitelaar, J. K., Richards, J. S., Hartman, C. A., Hoekstra, P. J., Franke, B., Heslenfeld, D. J., & Oosterlaan, J. (2017). Risk factors for comorbid oppositional defiant disorder in attention-deficit/hyperactivity disorder. European Child & Adolescent Psychiatry, 26(10), 1155–1164. https://doi.org/10.1007/s00787-017-0972-4

NSDUH. Substance Abuse and Mental Health Services Administration. (2019). Key substance use and mental health indicators in the United States: Results from the 2018 National Survey on Drug Use and Health (HHS Publication No. PEP19-5068, NSDUH Series H-54). Rockville, MD: Center for Behavioral Health Statistics and Quality, Substance Abuse and Mental Health Services Administration. Retrieved from https://www.samhsa.gov/data/

O’Connell, J., Gurdasani, D., Delaneau, O., Pirastu, N., Ulivi, S., Cocca, M., Traglia, M., Huang, J., Huffman, J. E., Rudan, I., McQuillan, R., Fraser, R. M., Campbell, H., Polasek, O., Asiki, G., Ekoru, K., Hayward, C., Wright, A. F., Vitart, V., … Marchini, J. (2014). A General Approach for Haplotype Phasing across the Full Spectrum of Relatedness. PLOS Genetics, 10(4), 1004234. https://doi.org/10.1371/journal.pgen.1004234

Oruc, I., Balas, B., & Landy, M. S. (2019). Face perception: A brief journey through recent discoveries and current directions. Vision Research, 157, 1–9. https://doi.org/10.1016/j.visres.2019.06.005

Polimanti, R., Agrawal, A., & Gelernter, J. (2017). Schizophrenia and substance use comorbidity: A genome-wide perspective. Genome Medicine, 9(1), 25. https://doi.org/10.1186/s13073-017-0423-3

Prom-Wormley, E. C., Ebejer, J., Dick, D. M., & Bowers, M. S. (2017). The genetic epidemiology of substance use disorder: A review. Drug and Alcohol Dependence, 180, 241–259. https://doi.org/10.1016/j.drugalcdep.2017.06.040

Sabia, S., Elbaz, A., Dugravot, A., Head, J., Shipley, M., Hagger-Johnson, G., Kivimaki, M., & Singh-Manoux, A. (2012). Impact of smoking on cognitive decline in early old age: The Whitehall II cohort study. Archives of General Psychiatry, 69(6), 627–635. https://doi.org/10.1001/archgenpsychiatry.2011.2016

Sabia, S., Marmot, M., Dufouil, C., & Singh-Manoux, A. (2008). Smoking history and cognitive function in middle age from the Whitehall II study. Archives of Internal Medicine, 168(11), 1165–1173. https://doi.org/10.1001/archinte.168.11.1165

Satterthwaite, T. D., Connolly, J. J., Ruparel, K., Calkins, M. E., Jackson, C., Elliott, M. A., Roalf, D. R., Hopson, R., Prabhakaran, K., Behr, M., Qiu, H., Mentch, F. D., Chiavacci, R., Sleiman, P. M. A., Gur, R. C., Hakonarson, H., & Gur, R. E. (2016). The Philadelphia Neurodevelopmental Cohort: A Publicly Available Resource for the Study of Normal and Abnormal Brain Development in Youth. NeuroImage, 124(0 0), 1115–1119. https://doi.org/10.1016/j.neuroimage.2015.03.056

Sayette, M. A. (2017a). The effects of alcohol on emotion in social drinkers. Behaviour research and therapy, 88, 76–89. https://doi.org/10.1016/j.brat.2016.06.005

Sun, F., Lei, Y., You, J., Li, C., Sun, L., Garza, J., Zhang, D., Guo, M., Scherer, P. E., Lodge, D., & Lu, X.-Y. (2019). Adiponectin modulates ventral tegmental area dopamine neuron activity and anxiety-related behavior through AdipoR1. Molecular Psychiatry, 24(1), 126–144. https://doi.org/10.1038/s41380-018-0102-9

Sun, G. Y., Xu, J., Jensen, M. D., & Simonyi, A. (2004a). Phospholipase A2 in the central nervous system: Implications for neurodegenerative diseases. Journal of Lipid Research, 45(2), 205–213. https://doi.org/10.1194/jlr.R300016-JLR200

Supek, F., Bošnjak, M., Škunca, N., & Šmuc, T. (2011). REVIGO Summarizes and Visualizes Long Lists of Gene Ontology Terms. PLOS ONE, 6(7), e21800. https://doi.org/10.1371/journal.pone.0021800

Swagerman, S. C., de Geus, E. J. C., Kan, K.-J., van Bergen, E., Nieuwboer, H. A., Koenis, M. M. G., Hulshoff Pol, H. E., Gur, R. E., Gur, R. C., & Boomsma, D. I. (2016). The Computerized Neurocognitive Battery: Validation, aging effects, and heritability across cognitive domains. Neuropsychology, 30(1), 53–64. https://doi.org/10.1037/neu0000248

Thapar, A., Cooper, M., Eyre, O., & Langley, K. (2013b). What have we learnt about the causes of ADHD? Journal of Child Psychology and Psychiatry, and Allied Disciplines, 54(1), 3–16. https://doi.org/10.1111/j.1469-7610.2012.02611.x

Trampush, J., Yang, M., Yu, J. et al. GWAS meta-analysis reveals novel loci and genetic correlates for general cognitive function: a report from the COGENT consortium. Mol Psychiatry 22, 336–345 (2017). https://doi.org/10.1038/mp.2016.244

Unterrainer, H. F., Hiebler-Ragger, M., Koschutnig, K., Fuchshuber, J., Ragger, K., Perchtold, C. M., Papousek, I., Weiss, E. M., & Fink, A. (2019). Brain Structure Alterations in Poly-Drug Use: Reduced Cortical Thickness and White Matter Impairments in Regions Associated With Affective, Cognitive, and Motor Functions. Frontiers in Psychiatry, 10. https://doi.org/10.3389/fpsyt.2019.00667

van Amsterdam, J., van der Velde, B., Schulte, M., & van den Brink, W. (2018). Causal Factors of Increased Smoking in ADHD: A Systematic Review. Substance Use & Misuse, 53(3), 432–445. https://doi.org/10.1080/10826084.2017.1334066

Wang, Yutong, Yao, L., & Zhao, X. (2020). Amygdala network in response to facial expression following neurofeedback training of emotion. Brain Imaging and Behavior, 14(3), 897–906. https://doi.org/10.1007/s11682-019-00052-4

WHO (2018), Global status report on alcohol and health 2018, available at https://www.who.int/substance_abuse/publications/global_alcohol_report/en/

Wickham, H. (2009). ggplot2: Elegant Graphics for Data Analysis. Springer-Verlag. https://doi.org/10.1007/978-0-387-98141-3

Winer, J. R., Maass, A., Pressman, P., Stiver, J., Schonhaut, D. R., Baker, S. L., Kramer, J., Rabinovici, G. D., & Jagust, W. J. (2018). Associations Between Tau, β-Amyloid, and Cognition in Parkinson Disease. JAMA Neurology, 75(2), 227–235. https://doi.org/10.1001/jamaneurol.2017.3713

Winterdahl, M., Noer, O., Orlowski, D. et al. Sucrose intake lowers μ-opioid and dopamine D2/3 receptor availability in porcine brain. Sci Rep 9, 16918 (2019). https://doi.org/10.1038/s41598-019-53430-9

Yates, A. D., Achuthan, P., Akanni, W., Allen, J., Allen, J., Alvarez-Jarreta, J., Amode, M. R., Armean, I. M., Azov, A. G., Bennett, R., Bhai, J., Billis, K., Boddu, S., Marugán, J. C., Cummins, C., Davidson, C., Dodiya, K., Fatima, R., Gall, A., … Flicek, P. (2020). Ensembl 2020. Nucleic Acids Research, 48(D1), D682–D688. https://doi.org/10.1093/nar/gkz966

Zhan, X., Hu, Y., Li, B., Abecasis, G. R., & Liu, D. J. (2016). RVTESTS: An efficient and comprehensive tool for rare variant association analysis using sequence data. Bioinformatics, 32(9), 1423–1426. https://doi.org/10.1093/bioinformatics/btw079

Zhou, H., Sealock, J. M., Sanchez-Roige, S., Clarke, T.-K., Levey, D. F., Cheng, Z., Li, B.,Polimanti, R., Kember, R. L., Smith, R. V., Thygesen, J. H., Morgan, M. Y., Atkinson, S. R., Thursz, M. R., Nyegaard, M., Mattheisen, M., Børglum, A. D., Johnson, E. C., Justice, A. C., … Gelernter, J. (2020). Genome-wide meta-analysis of problematic alcohol use in 435,563 individuals yields insights into biology and relationships with other traits. Nature Neuroscience, 23(7), 809–818. https://doi.org/10.1038/s41593-020-0643-5.

